# Clinical Evaluation of the PANDAA LASV RT-PCR Assay in Sierra Leone

**DOI:** 10.64898/2026.10.30.26366846

**Authors:** Donald S Grant, Caitlin R Thompson, John Sandi, Mariaam Manjia, Tiangay Kallon, Mohamed S. Kamara, Ibrahim Fofanah, Thomas Edwards, Tom E Fletcher, Rachel L Byrne

## Abstract

Molecular diagnosis of Lassa fever is complicated by extensive Lassa virus genetic diversity and limited laboratory capacity in endemic settings. The RealStar® Lassa Virus RT-PCR Kit 2.0 is a commonly used commercial dual-target assay for LASV detection, but its two-target format requires separate amplification reactions, increasing reagent use and workflow complexity. We present the diagnostic evaluation of the single-reaction, adaptive PANDAA LASV RT-PCR assay with the dual-target RealStar® Lassa Virus RT-PCR Kit 2.0 using 94 archived serum samples from Sierra Leone. 79 of these samples had valid paired results, and we report a positive percent agreement of 78.12% (95% CI 61.25%–88.98%), negative percent agreement of 93.62% (95% CI 82.84%–97.81%), and an overall agreement of 87.34% (95% CI 78.24%–92.98%); Cohen κ was 0.73 (95% CI 0.58–0.89). PANDAA showed substantial agreement and offers a potentially efficient single-well format, but further optimisation and prospective evaluation are needed.

## Introduction

Lassa fever is a viral haemorrhagic fever caused by Lassa Virus (LASV), an enveloped RNA virus of the family *Arenaviridae* and is endemic across several West African countries, including Nigeria, Sierra Leone, Liberia and Guinea [1].

Human infection occurs primarily through direct or indirect exposure to urine or faeces from infected rodents, including contamination of food and household surfaces, inhalation of aerosolised material during activities such as floor sweeping, and the handling or consumption of infected rodents [1,2]. Person-to-person transmission can also occur through direct contact with the blood or other bodily fluids of an infected individual. Although healthcare-associated transmission is particularly well documented and has resulted in outbreaks with substantial mortality among healthcare workers, transmission can also occur within households and communities following close contact with symptomatic cases [3]. Among hospitalised patients, case-fatality ratios of approximately 15–30% have been reported, although mortality varies substantially according to disease severity at presentation, case ascertainment and access to timely supportive care [4].

The true burden of Lassa fever remains poorly characterised. Historical estimates suggest that approximately 100,000–300,000 infections and around 5,000 deaths occur annually across West Africa [5], but these figures are associated with considerable uncertainty. Approximately 80% of LASV infections are thought to be asymptomatic or mild and are therefore unlikely to be identified through routine facility-based surveillance [6]. This under-ascertainment is compounded by limited access to laboratory confirmation, particularly outside established Lassa fever treatment centres and national or regional reference laboratories. Consequently, routinely reported cases are likely to represent only a fraction of infections occurring across endemic populations.

Rapid and accurate molecular diagnosis is therefore essential for Lassa fever. The early clinical presentation is non-specific and overlaps considerably with other causes of acute febrile illness [4,7], and early confirmation of LASV infection has important implications for clinical management, infection prevention and control, contact tracing and outbreak response. Reverse-transcription PCR (RT-PCR) is the principal molecular method used for confirmation of acute LASV infection. However, molecular diagnosis is complicated by the exceptionally high genetic diversity of LASV. At least seven major phylogenetic lineages have been described across West Africa, and nucleotide divergence between strains can be substantial (>27%) [8,9]. Sequence variation within primer- and probe-binding regions creates a particular challenge for conventional RT-PCR assays and can lead to lineage-dependent reductions in analytical sensitivity or failure to detect divergent strains [10,11]. No single conventional molecular assay has therefore provided uniformly optimal detection across the full genetic diversity of LASV.

The RealStar® Lassa Virus RT-PCR Kit 2.0 (altona Diagnostics, Germany) is one of the most established commercial molecular assays for LASV and is widely used by reference laboratories. The assay uses a dual-target strategy specifically intended to improve detection of genetically diverse LASV strains. It comprises two separate RT-qPCR assays, one targeting the glycoprotein precursor (GPC) gene on the S segment and the other targeting the L gene on the L segment. Consequently, testing a specimen against both LASV targets requires two RT-qPCR reactions per sample. The assay is CE-IVD marked and was among the assays evaluated in the WHO collaborative study used to establish the first WHO International Standard for LASV RNA [12] At present, there remains an important policy and product gap in that no LASV molecular diagnostic has been WHO prequalified or Emergency Use Listed, limiting the availability of an internationally recognised mechanism for assuring diagnostic quality and supporting procurement.

An alternative approach is the PANDAA LASV assay developed by Aldatu Biosciences (USA). PANDAA (Pan-Degenerate Amplification and Adaptation) uses an adaptive PCR methodology specifically designed to accommodate sequence polymorphisms within primer- and probe-binding regions and thereby enable broad detection across genetically diverse LASV lineages. PANDAA utilises ultra-degenerate primers with 3′ termini that adapt the probe binding site via site-directed mutagenesis to produce a conserved target sequence [13]. Unlike the RealStar® dual-assay approach, PANDAA contains the LASV target and internal control within a single reaction mix, allowing testing to be performed in one reaction per specimen. The LASV target is located within the L segment, with viral RNA detected using a FAM-labelled probe and the internal control detected using a VIC-labelled hydrolysis probe. Aldatu describes the assay as a pan-lineage LASV molecular diagnostic, with detection demonstrated across reference strains representing known LASV lineages. The assay is currently available for research use only.

Here, we compare the widely used dual-target RealStar® Lassa Virus RT-PCR Kit 2.0 to the single-reaction PANDAA LASV assay on stored serum samples.

## Methods

### Study population

Frozen (−80°C) serum samples from 94 individuals previously suspected of LASV infection were used for this retrospective clinical evaluation. Samples were collected from all suspected Lassa fever patients entering the Kenema Government hospital from 2^nd^ February 2023 to 5^th^ December 2025. Samples were identified for inclusion based on either the presence or absence of a previous diagnosis of Lassa Virus infection using the reference standard of RealStar® Lassa Virus RT-PCR Kit 2.0 performed at the time of sample collection. During this testing, 44 samples were classified as LASV positive and 50 as LASV negative by RealStar® 2.0. For the present study, stored samples were re-extracted, and each resulting RNA eluate was tested by both The RealStar® Lassa Virus RT-PCR Kit 2.0 and The PANDAA LASV RT-PCR kit. Upon re-extraction, 79 samples provided valid results with both assays and were included in the comparative assay analysis. Agreement between the two assays was subsequently assessed.

### Ethics

The study was approved by the Sierra Leone Ethics and Scientific Review Committee, Ministry of Health, Sierra Leone (SLESRC No. 002/07/2024), with Kenema Government Hospital as the approved study site. No patient-identifiable information was accessed by the analysis team

### Biosafety

Clinical samples were handled in a category 2+ laboratory in accordance with national biosafety guidelines. Extracted RNA was subsequently handled according to institutional molecular-diagnostic laboratory procedures.

### Sample extraction

Viral RNA isolation was performed using the QIAmp viral RNA mini kit (Qiagen, Germany) as per manufacturer’s instructions with a final elution volume of 60 µl.

### The RealStar® Lassa Virus RT-PCR

The RealStar® Lassa Virus RT-PCR Kit 2.0 was used as the comparator assay. The RT-PCR was performed on the Lightcycler 480 PCR system (Roche Diagnostics, Switzerland) following the manufacturer’s instructions for use (IFU). The RealStar® Lassa Virus RT-PCR Kit 2.0 detection kit targets the GPC gene and the L gene of Lassa Virus. Briefly, the protocol follows a reverse transcription step at 55 °C for 20 min, denaturation at 95 °C for 2 min, then 45 cycles of amplification at 95 °C for 15 s, 55 °C for 45 s, and 72 °C for 15 s, respectively. A volume of 10 µl of extracted RNA was used per reaction for a final volume of 30 µl. Results were interpreted in accordance with the assay instructions for use.

### The PANDAA LASV RT-PCR

The PANDAA LASV RT-PCR kit was used on the Lightcycler 480 PCR system following manufacturer’s instructions for use (IFU). The PANDAA LASV RT-PCR kit targets the L segment of the Lassa Virus. Briefly, the protocol follows a reverse transcription step at 50°C for 15 min, denaturation at 95 °C for 2 mins followed by 90°C for 3 s, followed by 10 cycles of the PANDAA adaption of 55°C for 30 s and 60 °C for 30 s, then 30 cycles of denaturation at 90 °C for 3 s and annealing/detection at 60 °C for 60 s. A volume of 10 µl of extracted RNA was used per reaction for a final volume of 20 µl. Amplification failure of the internal control in the absence of a positive LASV signal was reported as an invalid result. Results were classified as LASV positive, negative, or invalid according to the assay protocol.

### Statistical analysis

Statistical analyses were performed using R (R Foundation for Statistical Computing, Vienna, Austria). Agreement between the PANDAA LASV RT-PCR and RealStar® Lassa Virus RT-PCR assays was evaluated using a 2 × 2 contingency table. Positive percent agreement (PPA), negative percent agreement (NPA), positive predictive value (PPV), negative predictive value (NPV), likelihood ratios and overall percent agreement were calculated. PPV and NPV were estimated at an assumed disease prevalence of 46.8% based on the historical RealStar® Lassa Virus RT-PCR positivity proportion at the time of original sequential sample collection. The historical RealStar® results were used only to derive this prevalence estimate and were not included in the head-to-head agreement analysis. Ninety-five percent confidence intervals for proportions were estimated using the Wilson method. Agreement beyond chance was assessed using Cohen’s kappa coefficient with a 95% confidence interval. Differences between paired discordant results were assessed using a two-sided exact McNemar test.

PPA and NPA were calculated directly from the head-to-head comparison of the PANDAA LASV RT-PCR and RealStar® Lassa Virus RT-PCR following re-extraction of the stored serum samples. Disease prevalence was estimated from the historical RealStar® Lassa Virus RT-PCR results obtained at the time of original sequential sample collection. PPV and NPV were then calculated using the PPA and NPA derived from the head-to-head comparison together with the historical disease prevalence. A two-sided *p* value <0.05 was considered statistically significant.

## Results

Among the 79 samples with valid paired results, 25 samples were positive by both assays and 44 were negative by both assays. Ten samples produced discordance results between the two assays. Three were positive by PANDAA and negative by RealStar® and 7 were negative by PANDAA and positive by RealStar® (Table 1).

**Table 1.**
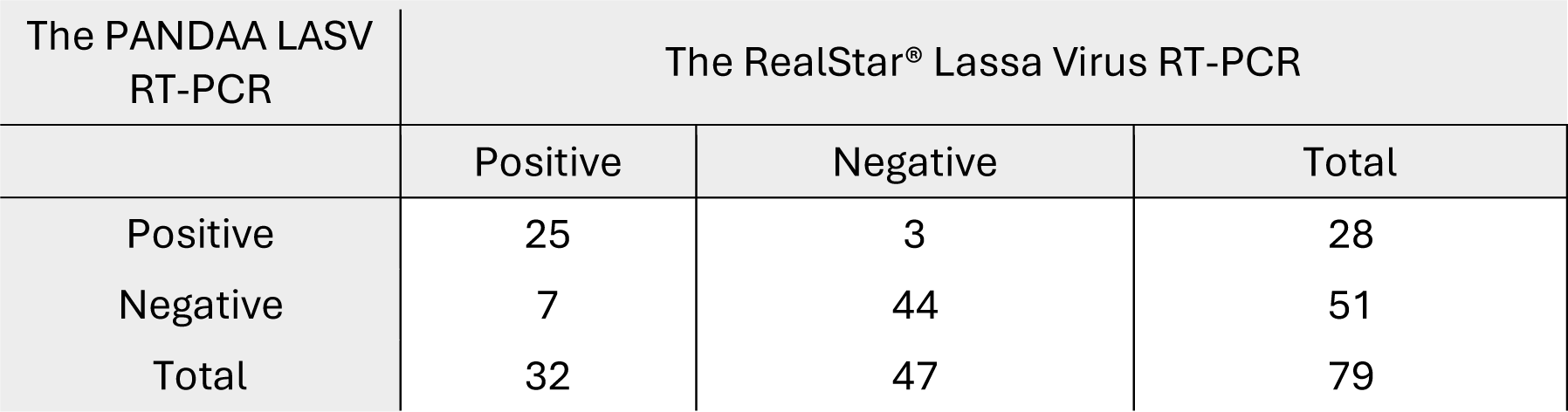
Results of the PANDAA LASV RT-PCR based on The RealStar® Lassa Virus RT-PCR. RT-PCR = Real-time polymerase chain reaction.

| The PANDAA LASV RT-PCR | The RealStar<sup>®</sup> Lassa Virus RT-PCR |  |  |
|  | Positive | Negative | Total |
|---|---|---|---|
| Positive | 25 | 3 | 28 |
| Negative | 7 | 44 | 51 |
| Total | 32 | 47 | 79 |

The RealStar® Lassa Virus RT-PCR was used as the comparator assay. The RealStar® Ct values were lower overall in the PANDAA positive samples than in the PANDAA negative samples (Figure 1). The median RealStar® Ct value using both S and L targets was Ct 22.52 in the PANDAA positive samples and Ct 31.19 in the PANDAA negative samples. RealStar® Ct values ranged from 19.34 to 33.42 in the PANDAA positive samples and from 27.70 to

**Figure 1.**
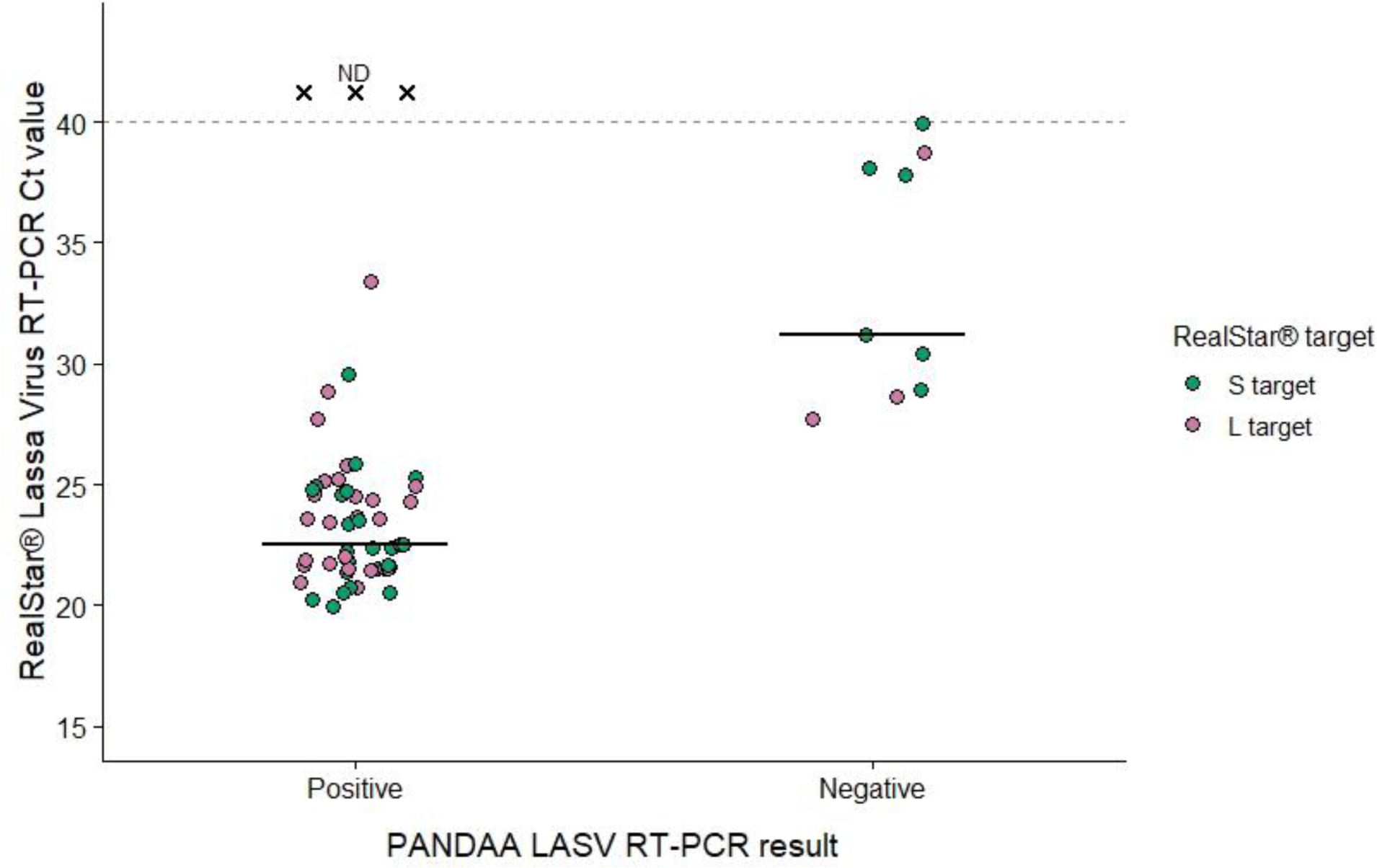
Distribution of RealStar® Lassa Virus RT-PCR Ct values according to PANDAA LASV RT-PCR result. PANDAA positive was defined as a Ct value below the cut-off (Ct 30) was obtained and PANDAA negative was defined as no Ct value. RealStar® S-target Ct values are shown as green circles and L-target Ct values as pink circles. Horizontal black lines show the median RealStar® Ct value. The dashed horizontal line denotes the RealStar® positivity threshold of Ct 40. Three PANDAA-positive samples that were negative by RealStar® RT-PCR (potential false positives) are represented by black crosses (×) labelled ND (not detected) above the Ct 40 threshold. Samples with no detectable RealStar® Ct values were excluded from the median calculations.

39.97 in the PANDAA negative samples.

The PPA of PANDAA was 78.12% (25/32; 95% CI 61.25–88.98%) and the NPA was 93.62% (44/47; 95% CI 82.84–97.81%). The disease prevalence was calculated as 46.8%, with a prevalence adjusted PPV of 91.50% (95% CI 82.17-100.00%) and a prevalence adjusted negative predictive value of 82.95% (95% CI 73.92-92.68%). The overall agreement between the two assays was 87.34% (95% CI 78.24 – 92.98%) with a Cohen’s kappa coefficient of 0.73 (95% CI 0.58 – 0.89). The positive likelihood ratio was 12.24 (95% CI 4.03-37.14) and negative likelihood ratio was 0.23 (95% CI 0.12 – 0.45) (Table 2). Comparison of the discordant paired results using a two-sided exact McNemar test showed no statistically significant difference between assays (p=0.344).

**Table 2.**
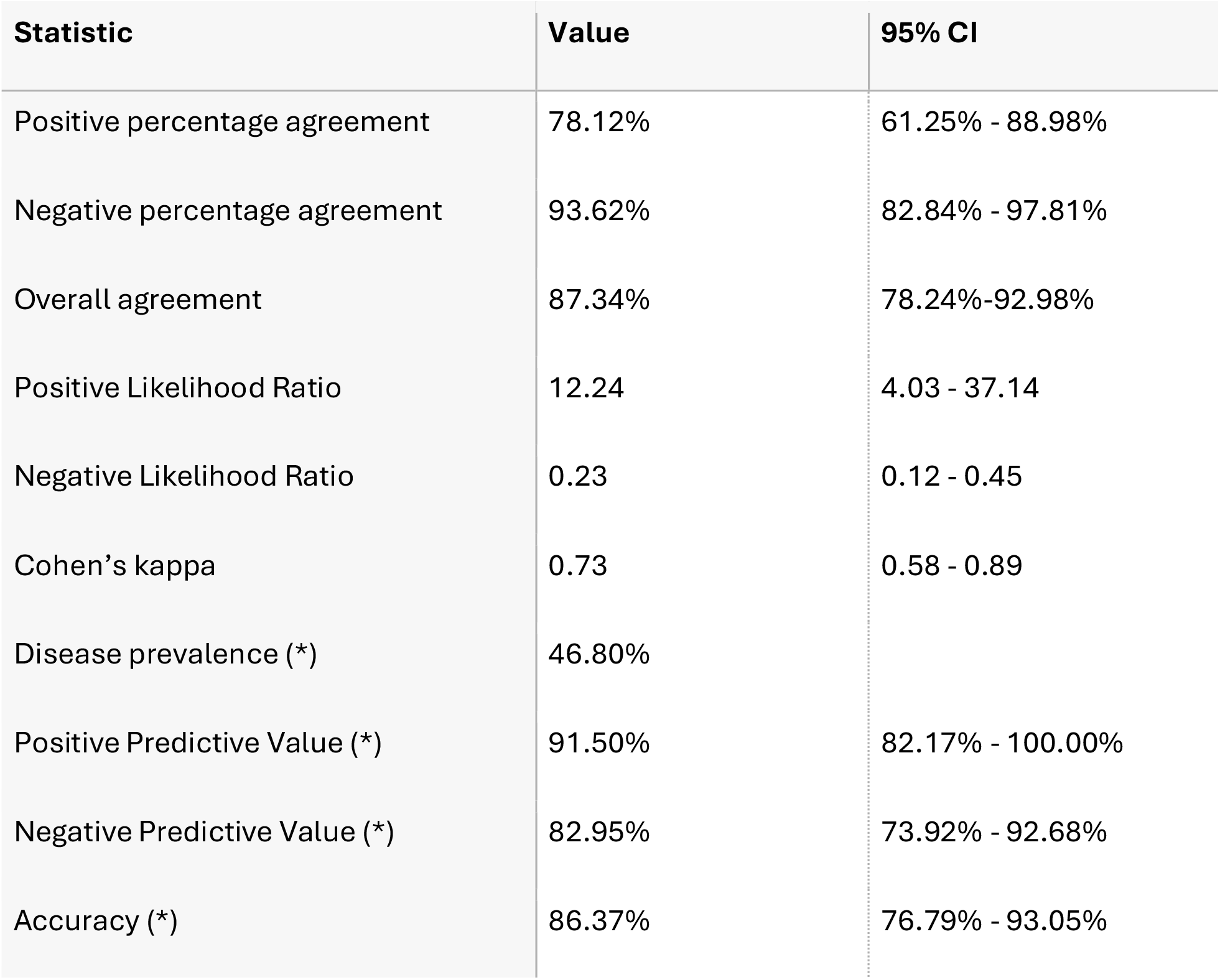
Agreement between PANDAA and RealStar® Lassa Virus RT-PCR assays. RT-PCR = Real-time polymerase chain reaction. CI = Confidence interval. (*) Positive predictive value, negative predictive value and accuracy calculated from historical RealStar® Lassa Virus RT-PCR positivity proportion at the time of original sequential sample collection.

| **Statistic** | **Value** | **95% CI** |
|---|---|---|
| Positive percentage agreement | 78.12% | 61.25% - 88.98% |
| Negative percentage agreement | 93.62% | 82.84% - 97.81% |
| Overall agreement | 87.34% | 78.24%-92.98% |
| Positive Likelihood Ratio | 12.24 | 4.03 - 37.14 |
| Negative Likelihood Ratio | 0.23 | 0.12 - 0.45 |
| Cohen’s kappa | 0.73 | 0.58 - 0.89 |
| Disease prevalence (*) | 46.80% |  |
| Positive Predictive Value (*) | 91.50% | 82.17% - 100.00% |
| Negative Predictive Value (*) | 82.95% | 73.92% - 92.68% |
| Accuracy (*) | 86.37% | 76.79% - 93.05% |

## Discussion

Rapid and reliable molecular diagnostics for Lassa fever remain challenging, particularly in endemic settings where confirmatory testing is still concentrated largely within regional or national reference laboratories, while delays in sample transport and limitations in laboratory infrastructure can cause delays in diagnosis. In addition, the substantial genetic diversity of LASV can have an impact on the performance of a molecular assay by FIND (Foundation for Innovative New Diagnostics) target product profile [14] for confirmatory Lassa fever diagnostics specifies broad inclusivity across LASV lineages I-VII together with operational characteristics suitable for use in reference laboratories and, optimally, district and regional hospital laboratories. Therefore, the development and evaluation of molecular assays such as PANDAA should aim to maintain broad target detection, ideally with a simplified testing workflow.

This study evaluated the research-use-only PANDAA LASV RT-PCR assay against the RealStar® Lassa Virus RT-PCR Kit 2.0 using retrospective stored serum samples from the Kenema Government Hospital, Sierra Leone.

Among the 79 samples that generated valid results with both assays, PANDAA had an overall percent agreement with RealStar® of 87.34% (95% CI 78.24%–92.98%) and a Cohen κ of 0.73 (95% CI 0.58–0.89), indicating substantial agreement beyond chance. NPA was higher than PPA at 93.62% (95% CI 82.84%–97.81%) compared with 78.12% (95% CI 61.25%–88.98%), respectively. Ten samples produced discordant valid results, comprising 3 PANDAA-positive/RealStar®-negative and 7 PANDAA-negative/RealStar®-positive samples (S. Table 1). The difference between discordant results was not statistically significant y exact McNemar testing (*p*=0.344), although the small number of discordant results reduces the precision of this comparison. RealStar® Ct values were generally higher among PANDAA-negative samples (Figure 1) which suggests that reduced detection by PANDAA may be more likely in samples with lower target concentrations compared to the RealStar® assay. However, the sample size was relatively small (n = 32) and confirmation of this trend will require a larger prospective sample evaluation.

No single reference test currently provides a definitive determination of Lassa fever infection status [15]. However, the FIND TPP for confirmatory Lassa diagnostics specifies diagnostic performance comparison to RealStar® Lassa Virus 2.0 RT-PCR assay (Altona, Germany) with a minimum diagnostic sensitivity and specificity targets of ≥95.00% and optimal targets of ≥98.00% [14]. As the comparison with RealStar® does not assume that every RealStar® result represents the true infection status, we have retained the use of PPA and NPA rather than sensitivity and specificity.

The PANDAA PPA of 78.12% and NPA of 93.62% were below the minimum TPP thresholds (≥95.00%). However, the NPA of 93.62% had a 95% confidence interval of 82.84–97.81%, which includes values above the TPP threshold, indicating substantial uncertainty around the point estimate. However, the PPA confidence interval (61.25–88.98%) remained below the TPP target, suggesting that further evaluation and/or optimisation of this assay will likely be required. This comparison is useful for preliminary assessment of the assay but should not be interpreted as a definitive estimate of clinical sensitivity or specificity.

Despite these performance limitations, the PANDAA format has potential operational advantages. PANDAA detects the LASV target and internal control within a single reaction well, whereas RealStar® uses separate reactions for its S- and L-segment LASV targets. The single-reaction format could therefore reduce reagent use and thermocycler capacity required per specimen, potentially increasing throughput during periods of high testing demand. Additionally, the PANDAA assay has been designed to tolerate sequence variation within the primer-probe-binding regions through the use of degenerate primers and adaptation of the probe-binding site during amplification, potentially reducing susceptibility to target-site polymorphisms [13]. Conventional RT-PCR assays can be affected by sequence variation, although the dual-target design of RealStar® may reduce this risk by providing redundancy if one target is affected. Sequencing was performed on a small subset of samples, all of which were identified as belonging to LASV clade IV. However, further sequencing analysis of all samples is planned to assess genetic diversity and investigate whether sequence variation may have influenced assay performance. Beyond analytical performance, operational characteristics are also important, as identified by the FIND TPP, which emphasizes time to result and the ability to test multiple samples simultaneously [14].

This study has several limitations. The specimen panel was retrospective and selected according to historical RealStar® status rather than through consecutive enrolment and therefore does not represent disease prevalence in the source population. There was no independent reference assay available to resolve discordant PANDAA and RealStar® results. The relatively small number of comparator-positive samples resulted in wide confidence intervals around PPA, and viral lineage was determined for only a subset of samples, and all sequenced viruses belonged to clade IV. As the LASV genetic diversity varies geographically [16], the performance of both RT-PCR assays may vary between circulating lineages and the sequence variation within the assay target regions. Prospective evalua tion using consecutively collected samples, characterized viral lineages and viral loads, and an independent method for resolving discordant results would provide a more definitive assessment of clinical performance.

Overall, PANDAA demonstrated substantial agreement with RealStar® and particularly strong negative agreement. Although the observed PPA and NPA did not currently meet the minimum FIND TPP. A large prospective evaluation using freshly collected or minimally freeze–thawed samples and representation of multiple LASV lineages and viral loads, is needed. These further studies will be important to determine whether the operational advantages of PANDAA’s single-reaction, adaptive PCR format can be combined with the diagnostic performance required for broader use in Lassa-endemic settings.

## Data Availability

All data produced in the present study are available upon reasonable request to the authors

## Supplementary

**S. Table 1.**
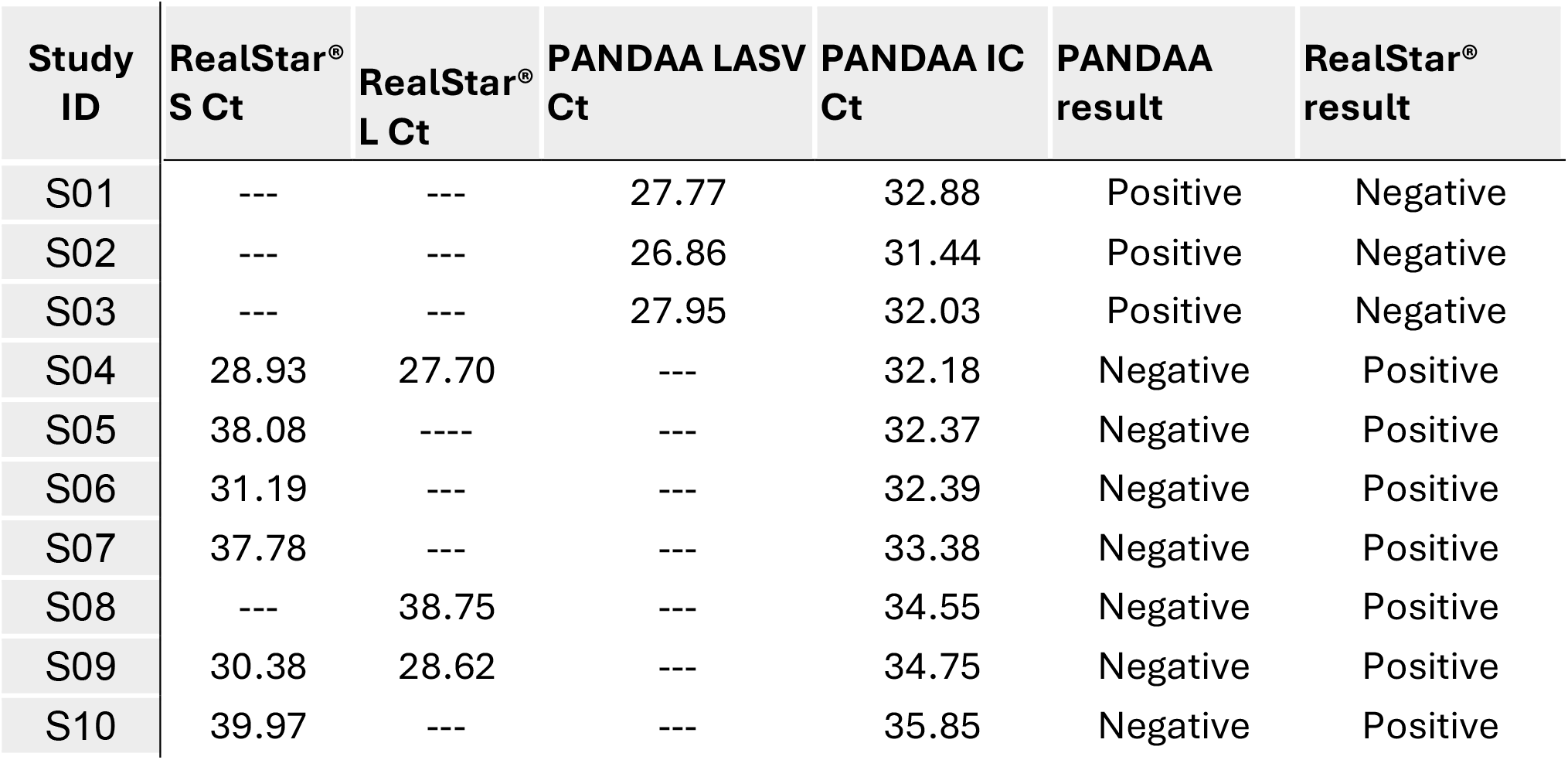
Discordant PANDAA and repeat RealStar® results.

| **Study ID** | **RealStar<sup>®</sup> S Ct** | **RealStar<sup>®</sup> L Ct** | **PANDAA LASV Ct** | **PANDAA IC Ct** | **PANDAA result** | **RealStar<sup>®</sup> result** |
|---|---|---|---|---|---|---|
| S01 | --- | --- | 27.77 | 32.88 | Positive | Negative |
| S02 | --- | --- | 26.86 | 31.44 | Positive | Negative |
| S03 | --- | --- | 27.95 | 32.03 | Positive | Negative |
| S04 | 28.93 | 27.70 | --- | 32.18 | Negative | Positive |
| S05 | 38.08 | ---- | --- | 32.37 | Negative | Positive |
| S06 | 31.19 | --- | --- | 32.39 | Negative | Positive |
| S07 | 37.78 | --- | --- | 33.38 | Negative | Positive |
| S08 | --- | 38.75 | --- | 34.55 | Negative | Positive |
| S09 | 30.38 | 28.62 | --- | 34.75 | Negative | Positive |
| S10 | 39.97 | --- | --- | 35.85 | Negative | Positive |

## Notes

### Competing Interest Statement

The authors have declared no competing interest.

### Summary of Updates

Correct name of PANDAA RT-PCR kit

### Funding

- Wellcome Trust

